# Innovation of Audio-Visual Triage system in Combating the Spread of COVID-19 Infection and its efficacy: A Novel Strategy

**DOI:** 10.1101/2020.11.06.20223040

**Authors:** Muhammad Mansoor Hafeez, Mohammad Azhar, Hafiz Rizwan Zafar Chudhary, Muhammad Asim Rana, Arif Malik

## Abstract

During the novel coronavirus pandemic, also known as SARS-CoV-2 or COVID-19 pandemic, frontline healthcare professionals suffered psychological as well as pathological trauma due to the lack of preparation to cope with this unforeseen situation. The protocols to prevent the spread of this disease proved to be less effective than anticipated. In these circumstances, improvement of the existing triage system was felt and an AUDIO-VISUAL TRIAGE (AVT) system was introduced to enhance confidence as well as increase the safety of frontline healthcare professionals. The current analysis was performed from March 21, 2020, to April 28, 2020, until the completion of sixty response forms, at Bahria Town International Hospital, Lahore. Thirty participants (Group A) deployed on visual triage and other thirty (Group B) on AVT for screening suspected cases of COVID-19 infection. Anxiety levels were measured by using the GAD-7 scoring system and the participants of both groups were periodically tested for COVID-19 infection by PCR. Independent t-test was used to evaluate the significance of different variables at a confidence level of 95%. The result of the current study revealed the effectiveness of AVT for the screening of COVID-19 patients. There was a statistically significant increase in anxiety levels and severe acute respiratory syndrome coronavirus 2 (SARS-CoV-2) infection rate in group A as compared to group B. Almost all participants in group A wanted to shift their place of work or ready to quit the job if they were forced to perform their duties at the same visual triage. AVT system for COVID-19 screening found to be more safe and less stressful than visual triage. It is not only a simple and effective way to prevent the spread of diseases but also boosted the confidence of frontline healthcare professionals to fight against coronavirus spread.

## INTRODUCTION

The pandemics are frightening for everyone, especially for healthcare workers. The most important aspect of dealing with the pandemic is to protect and save the healthcare workers so that they can treat the rest of the people effectively (Mason et al; 2020). Triage is the forefront of an emergency department of an acute hospital which becomes more fundamental during the pandemics. keeping in view the COVID-19 prevailing situation, a new Triage system to filter and classify the patients reaching hospital with suspected corona virus infection was developed, named as “AUDIO-VISUAL TRIAGE” system abbreviated as AVT. The recent alarming situation of corona virus infection that has spread to 196 countries around the globe and resulted in failure of routine triage of Emergency Department (Habibi et al; 2020).

### Visual Triage System

From the beginning of rapid increase in coronavirus cases every hospital in Pakistan placed a screening triage named as visual triage system outside the main entry door. So a two-stage triage is introduced, first a visual triage (outside at entry door) & a routine triage (inside Emergency Department). But at visual triage, healthcare providers felt scared and threatened due to proximity with the patients and their attendants. As a result, they became reluctant and even some of them refused to perform duties at visual triage desk.

### Audio-Visual Triage System

Keeping this in mind, an Audio System was added to visual triage that resulted in an AVT. Protocols of AVT included, a medical personal sits on TRIAGE DESK (with glass barrier sheet on it) wearing personal protective equipment (PPE) at a distance of more than 6 feet from the PATIENT DESK. Both desks are connected with Non-touchable MIC SYSTEM for communication (fig 1,2). An Assessment Form with the scoring system is filled by the triage medical staff member and further management of the patient is based on the score achieved (Sivan et al; 2020). In doubtful cases, to reduce the chance of wrong decision (which can be devastating for the whole hospital), an Emergency Doctor re-assess the case and decides accordingly. Then Pathways for the patient management is developed according to the available facilities of the respective hospital and the instructions of Government. The suspected or confirmed cases are either treated in Isolation wards or Referred to respective Centre (designated by The Government) with all the facilities.

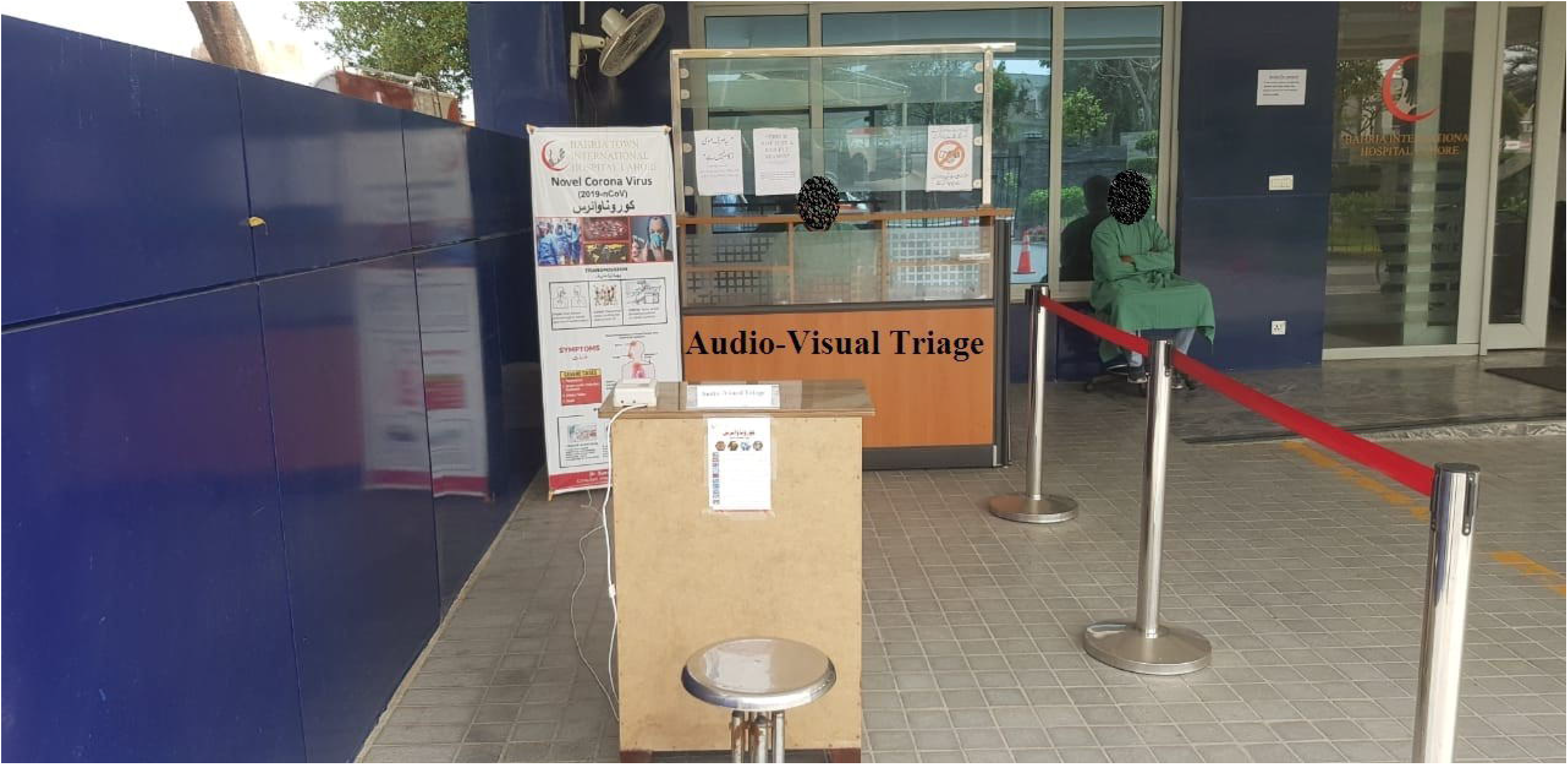

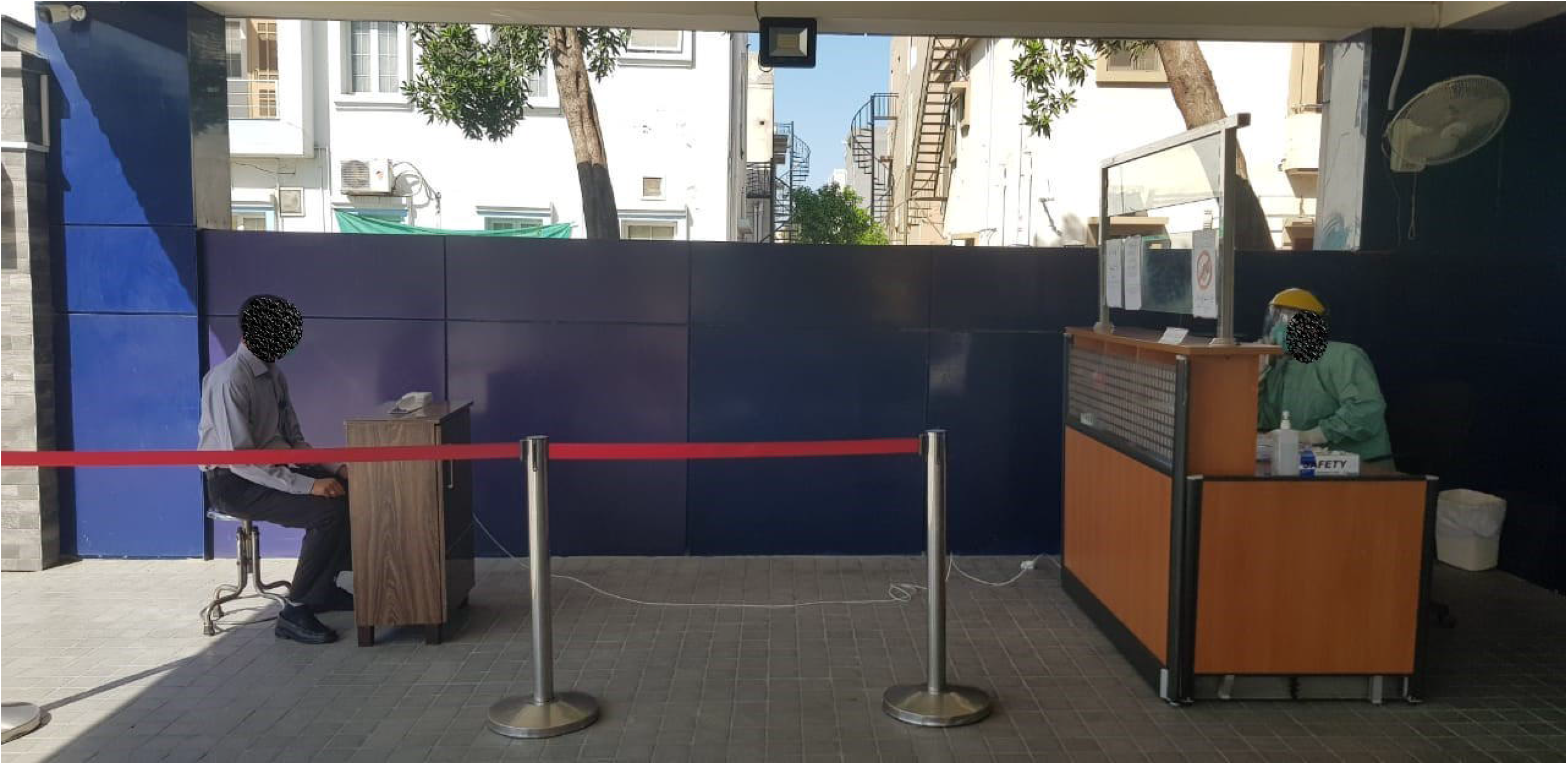

## MATERIAL AND METHODS

A total of sixty (n=60) participants was recruited to study the effectiveness of AVT System, in the current cross-sectional comparative study. They were further divided into two groups. Participants of group A were performing their duty on visual triage while those of group B were on AVT system for screening the COVID-19 suspected patients. Participants of both groups were provided with the same quality of PPEs. Those participants having previous psychological illness, frequently used to use the washroom, had non-serious attitude and poor compliance with the protocols of wearing PPEs were excluded from the study. After a week every participant was tested by PCR for COVID-19 infection. Questionnaires were distributed among the participants and the response form was collected for further analysis. The study was approved by the ethical review committee of Bahria Town International Hospital Lahore (31.5204° N; 74.3587° E), Pakistan. For the assessment of anxiety, GAD-7 scoring system was used and outcomes were assessed according to the Spitzer et al., 2006 scoring system. The GAD-7 score is calculated by assigning scores of 0, 1, 2, and 3, to the response categories of ‘not at all’, ‘several days’, ‘more than half the days’, and ‘nearly every day’, respectively, and adding together the scores for the seven questions. Scores of 5, 10, and 15 are taken as the cut-off points for mild, moderate and severe anxiety, respectively. Using the threshold score of 10, the GAD-7 has a sensitivity of 89% and a specificity of 82% for GAD. It is moderately good at screening three other common anxiety disorders - panic disorder (sensitivity 74%, specificity 81%), social anxiety disorder (sensitivity 72%, specificity 80%) and post-traumatic stress disorder (sensitivity 66%, specificity 81%)

Apart from the GAD-7 scoring, another question, related to job satisfaction, was asked. The data were statistically analyzed with the help of independent t-test using SPSS Software (version 22). In addition, the *p*-values of less than 0.05 remained statistically significant.

## RESULTS

The result of the current study revealed the effectiveness of AVT for the screening of COVID-19 patients. Total of sixty (n=60) participants were allocated equally in two groups. Group A were performing their duties on visual triage with male to female ratio was 21:9 (mean age 28.3 ± 4.01) whereas group B were indulged in screening COVID-19 patients on Audio-Visual Triage (male to female ratio 22:8, mean age=26.56 years). (table-1). The significantly increased levels of anxiety in group A were found as compared to group B (*p*=0.025).

**Table 1.**
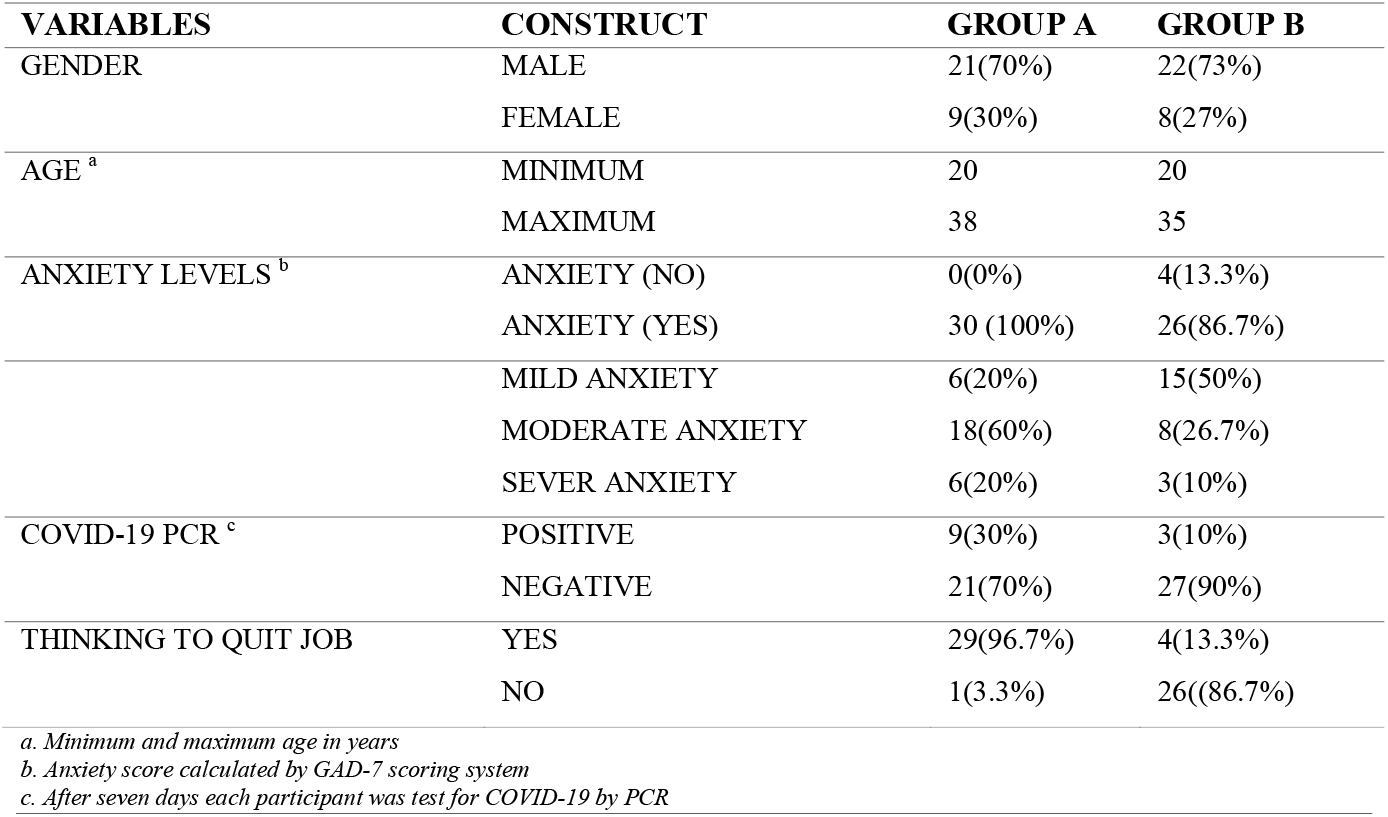
Distribution of Demographic and Other Variables Used for The Assessment of The Audio-Visual Triage System.

| VARIABLES | CONSTRUCT | GROUP A | GROUP B |
| --- | --- | --- | --- |
| GENDER | MALE | 21(70%) | 22(73%) |
|  | FEMALE | 9(30%) | 8(27%) |
| AGE <sup>a</sup> | MINIMUM | 20 | 20 |
|  | MAXIMUM | 38 | 35 |
| ANXIETY LEVELS <sup>b</sup> | ANXIETY (NO) | 0(0%) | 4(13.3%) |
|  | ANXIETY (YES) | 30 (100%) | 26(86.7%) |
|  | MILD ANXIETY | 6(20%) | 15(50%) |
|  | MODERATE ANXIETY | 18(60%) | 8(26.7%) |
|  | SEVER ANXIETY | 6(20%) | 3(10%) |
| COVID-19 PCR <sup>c</sup> | POSITIVE | 9(30%) | 3(10%) |
|  | NEGATIVE | 21(70%) | 27(90%) |
| THINKING TO QUIT JOB | YES | 29(96.7%) | 4(13.3%) |
|  | NO | 1(3.3%) | 26((86.7%) |

*a. Minimum and maximum age in years*
*b. Anxiety score calculated by GAD-7 scoring system*
*c. After seven days each participant was test for COVID-19 by PCR*

At the end of the study, there were more COVID-19 PCR positive participants in group A (*p*=0.001). (graph-1). Almost all participants in group A want to shift their place or ready to quit the job if they were forced to perform the duties at the same visual triage (*p*=0.004). (table-2)

**Table-2:** Outcomes of Different Variables for Assessment of AVT* System.

| VARIABLES | <sup>1</sup>GROUP A | <sup>2</sup>GROUP B | P-VALUE |
| --- | --- | --- | --- |
| ANXIETY SCORE | <sup>a</sup>$3 \pm 0.117$ | $2.33 \pm 0.154$ | $0.025^d$ |
| POSITIVE COVID-19 PCR | <sup>b</sup>$1.30 \pm 0.085$ | $1.10 \pm 0.056$ | $0.001^d$ |
| DESIRE TO QUIT JOB | <sup>c</sup>$1.97 \pm 0.033$ | $1.13 \pm 0.063$ | $0.004^d$ |

*a. Mild Anxiety coded as 1, moderate as 2 and sever as 3*
*b. COVID-19 Positive PCR coded as 2 and Negative as 1*
*c. Answer NO coded as 1 and YES as 2*
*d. Independent t-test to find out the significance of variance*
*1. Thirty participant performing their duties at visual triage screening system*
*2. Thirty participant performing their duties at AVT system*
*\* AUDIO-VISUAL TRIAGE*

**Graph-1:**
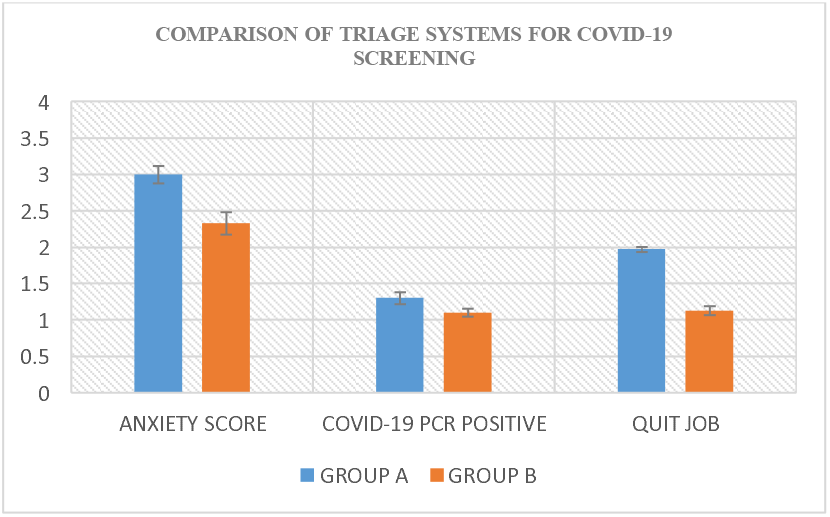
Comparison of Triage Systems for COVID-19 Screening.

## DISCUSSION

Triage is an important area of an Emergency Department of any acute Hospital. In this process, patients are classified according to the severity of their diseases so that the acutely sick and more serious patients are seen before the less serious (Bruijns et ak; 2008). Normally, the severity of the disease is assessed in a Triage Room of hospital where the most senior nurse asks about the presenting complaints of patients and checks the vitals of patients before allocating any Category / Class to them. Most of the triage systems have five Categories or Classes. Patients of Category I are the most serious patients, to be seen straight away and patients of Category V are less urgent and would be seen at the last. There are few Triage systems in the world e.g.; Manchester Triage, Australian Triage and Canadian Triage (FitzGerald et al; 2020). But these Triage systems are useful only for classifying patients having routine diseases but cannot be used during disasters like COVID-19 pandemic (Bazyar et al; 2020). If traditional triage criteria are used in pandemics, it can result in the contamination of the Emergency Department and ultimately the whole hospital which may need to be fumigated. Moreover, the Triage staff is liable to be infected and feel threatened which can result in a sensation of frustration and decrease job satisfaction (Forsgärde et al; 2013). An AVT proved to be a simple and effective solution to minimize the elevated psychological stress and more effective triage system in prevention of SARS-CoV-2 infection. Moreover, the pandemics do not give much time for preparation (by the Medical & nonmedical teams) and require big budgets as well as resources by the government. There is a need of increase number of healthcare workers, their training to deal with the pandemic disease, availability of big isolations areas, equipment, accessories and logistics in a short time (Kumar et al; 2020). These become more difficult to manage in poor economic countries. But by taking simple and cost-effective measures like AVT system, for screening suspected subjects, can boost the combating capabilities of frontline healthcare professionals against SARS-CoV-2 virus spread.

## CONCLUSION

The AVT system is more safe and effective strategy to prevent the spread of COVID-19 infection as patients are directed to their respective areas, without compromising the management. So it can be concluded AVT system should be recommended for all the hospitals dealing with suspected cases of Corona virus infection as it has been found practical, safe and confidence booster for frontline medical fighters on the basis of results of the current study.

## LIMITATIONS

The residence of participants located at different places like hostels or homes. There would be a chance of virus transmission in participants form their residence.

## Data Availability

The Data will be provided on demand

